# Awareness, Experiences, and Attitudes Toward Preprints Among Medical Academics

**DOI:** 10.1101/2025.04.09.25325524

**Authors:** Mustafa Sevim, Burak Karameşe, Zafer Alparslan

## Abstract

**Background:** Preprints—scientific manuscripts shared publicly prior to formal peer review—are gaining momentum across academic disciplines. However, their adoption in clinical and biomedical sciences remains limited, particularly in countries where traditional publishing norms prevail. Editorial ambiguity and a lack of national policy further complicate their use.

**Objective:** This study aimed to assess the awareness, experiences, and attitudes of medical academics at a major university in İstanbul, Türkiye toward preprints, and to explore the editorial landscape through both journal editor feedback and a review of journal-level preprint policies.

**Methods:** A cross-sectional survey was conducted with 103 medical faculty members. The questionnaire included demographic questions, Likert-scale and multiple-choice items assessing knowledge, familiarity, and attitudes toward preprints, as well as open-ended items to explore concerns. A “Preprint Test Score” (0–4) was developed to quantify objective knowledge. Subgroup analyses were conducted by age (<40 vs. ≥40 years) and academic discipline (basic vs. clinical sciences). Additionally, five journal editors responded to open-ended questions about preprint policies, and 118 biomedical journals were manually reviewed for their declared stances on preprints and article processing charges (APCs).

**Results:** Only 42.9% of participants reported familiarity with the concept of preprints, and 13% had previously published on a preprint server. Misconceptions about ethics, peer review, and compatibility with journal policies were common. Subgroup analysis revealed that older participants scored higher on the Preprint Test and had more experience with preprint publishing. Further younger academics expressed less openness toward future use. Clinical faculty were generally more hesitant than basic science faculty, although both groups raised concerns about the academic recognition of preprints. Editorial responses reflected a mix of cautious endorsement and scepticism. Among the 118 biomedical journals reviewed, most lacked clear preprint policies, while a small number either explicitly prohibited or permitted them.

**Conclusion:** There is limited awareness and cautious engagement with preprints among medical academics and editors in Türkiye. Generational and disciplinary differences further influence knowledge and attitudes. The lack of clear editorial guidance from biomedical journals may reinforce academic uncertainty. Tailored educational initiatives, transparent journal policies, and institutional support will be essential to foster a more open and inclusive scientific publishing environment.

## INTRODUCTION

Preprints—manuscripts publicly shared prior to peer review—have transformed the pace and openness of scholarly communication. Widely adopted in fields such as physics and computer science for decades, preprints enable rapid dissemination, open peer commentary, and broader visibility of research findings(1). In recent years, the biomedical community has increasingly engaged with preprint platforms, particularly during public health crises such as the COVID-19 pandemic. For instance, between June 2020 and June 2022, the U.S. National Library of Medicine made more than 3,300 NIH-funded COVID-19 preprints accessible in PubMed Central (PMC), marking a pivotal shift toward preprint integration in mainstream biomedical publishing (2).

Despite this global momentum, the adoption of preprints in clinical and medical sciences remains uneven(3). In countries like Türkiye, where academic evaluation systems and journal structures still emphasize traditional peer-reviewed publication, the concept and utility of preprints are often misunderstood or undervalued. Anecdotal observations suggest hesitancy among medical faculty, fuelled by concerns about plagiarism, duplication, and lack of recognition in academic promotion criteria.

Journal editors also play a crucial role in shaping scholarly norms. Editorial policies on preprints vary widely across journals: while some encourage their use, others either prohibit them or do not explicitly mention them at all (4–7). The absence of a clear preprint policy creates uncertainty for authors and may contribute to low adoption, particularly among early-career researchers concerned about publication eligibility (8).

While international studies have explored general attitudes toward preprints (3,9–11), little is known about how these perspectives vary within academic subgroups. Factors such as career stage and departmental discipline may influence both knowledge and perception. For instance, basic science researchers are often more open to experimentation with publishing models, whereas clinical academics may prioritize peer-reviewed evidence with clear implications for practice (12). Similarly, younger faculty may view preprints as tools for early visibility and career advancement, while more senior academics may adhere to traditional notions of scholarly validation and prestige.

To date, no systematic assessment has examined the knowledge, attitudes, and editorial perspectives on preprints within Türkiye’s medical academic community. This study seeks to address that gap through a mixed-method approach that combines: [1] a structured survey conducted at a major medical university in İstanbul, [2] qualitative feedback from editors of Turkish biomedical journals, and [3] a comprehensive review of journal-level preprint and APC policies. In addition to characterizing general patterns, we examine subgroup differences by age and academic discipline to uncover nuanced barriers and opportunities for preprint adoption in the evolving landscape of scientific communication.

## MATERIALS AND METHODS

### 1. Study Design and Setting

This study employed a cross-sectional survey design conducted at a major medical university in İstanbul (Ethics Committee approval number: 09.2024.600). The goal was to evaluate the awareness, knowledge, and attitudes of medical academics toward preprints, and to explore perceived barriers to preprint use. All graphs Created in https://BioRender.com

### 2. Participant Recruitment and Data Collection

A structured online questionnaire was distributed via institutional email lists and internal professional networks. The survey targeted medical academics from a variety of departments and academic ranks at a major medical university in İstanbul. No personal identifiers were collected, and all responses were anonymized and aggregated for analysis.

Although the total number of participants was 108, not all respondents answered every question. Some skipped certain items, particularly in the later sections of the survey. As a result, the number of responses varies across different variables, and this is reflected in the sample sizes reported in the Results section.

### 3. Survey Instrument

The questionnaire included, demographic items (e.g., age, academic title, department), multiple-choice and likert-scale questions assessing familiarity with preprints, previous use, attitudes toward preprints and peer review, and expectations for scientific quality and open-ended questions capturing perceived barriers and concerns related to preprint use.

To quantify objective knowledge of preprints, a “Preprint Test Score” was generated from four multiple-choice questions. Participants received one point for each correct response, resulting in a total score ranging from 0 to 4.

### 4. Subgroup Analyses

To explore differences in preprint engagement and perceptions, participants were stratified into subgroups based on:

- **Age:** younger than 40 years vs. 40 years or older,
- **Academic discipline:** basic sciences (e.g., physiology, microbiology) vs. clinical sciences (e.g., internal medicine, surgery).

Subgroup comparisons were made for knowledge scores, attitudes, and future intent to use preprints, allowing identification of generational and disciplinary trends.

### 5. Editorial Perspectives

To gather complementary insights into institutional attitudes toward preprints, we systematically reviewed all journals indexed in the Web of Science (InCites Dataset + Emerging Sources Citation Index), filtered for Türkiye as the country of publication and covering the time period from 2019 to 2023. From an initial list of 280 journals, 264 remained after excluding duplicates, inaccessible websites, and journals with unclear policies.

These journals were manually categorized into biomedical and non-biomedical fields based on their scope and published content. Journals within the disciplines of medical sciences, pharmacology, biology, veterinary sciences, and nursing were classified as biomedical. Based on this classification, we identified 118 biomedical journals indexed in the dataset and based in Türkiye (as of April 2025).

Editors of these journals were contacted via email and invited to respond to three open-ended questions:

- Their journal’s current stance on preprints,
- The rationale behind that stance,
- Their views on the future role of preprints in academic publishing.

A total of seven editors responded with usable feedback. Given the limited number of responses, the data were summarized descriptively rather than subjected to formal thematic analysis.

### 6. Journal Policy Review

In parallel, the same 118 biomedical journals were reviewed to assess their formal policies on preprints and Article Processing Charges (APCs). Policy information was manually collected by examining publicly available sections of the journals’ websites, including “Instructions for Authors,” “Editorial Policy,” and “Ethical Guidelines.”

This assessment was conducted in two rounds—first in February 2024 and again in April 2025— to track any changes in policy over time.

Journals were categorized as follows:

- **Preprint Policy**: *Allowed, Prohibited*, or *Not Mentioned*;
- **APC Policy**: *Obligatory, Free*, or *Case-to-case* (where charges depend on article type or other conditions).

This analysis provided insight into the editorial infrastructure surrounding preprints in Türkiye and helped contextualize how journal-level policies may influence researchers’ behaviour and perceptions.

## RESULTS

### 1. Preprint and APC Policies of Turkish Biomedical Journals

As part of our systematic review of biomedical journal policies in Türkiye, we analysed the trends in preprint and Article Processing Charge (APC) policies at two time points: February 2024 and April 2025. This dual-timepoint approach aimed to assess how Turkish biomedical journals are evolving in response to global shifts in open science practices and publication economics.

### 1.1. Preprint Policies

Preprint policies were categorized into three groups:

- **Allowed**: Journals explicitly welcome or permit submissions that were previously posted as preprints.
- **Prohibited**: Journals clearly disallow submissions that have appeared as preprints.
- **Not Mentioned**: No reference to preprints could be found on the journal’s official website.

Although the majority of journals still do not provide explicit guidance on preprints, our comparison over time revealed a modest but meaningful shift toward acceptance. Between February 2024 and April 2025, the number of journals explicitly allowing preprints increased from 27 to 33, while those with no policy decreased from 89 to 82. One additional journal began explicitly prohibiting preprints, increasing that category from two to three journals. These findings suggest a gradual trend toward policy transparency and a slow but positive normalization of preprint culture among Turkish biomedical journals (**Figure 1A**).

**Figure 1.**
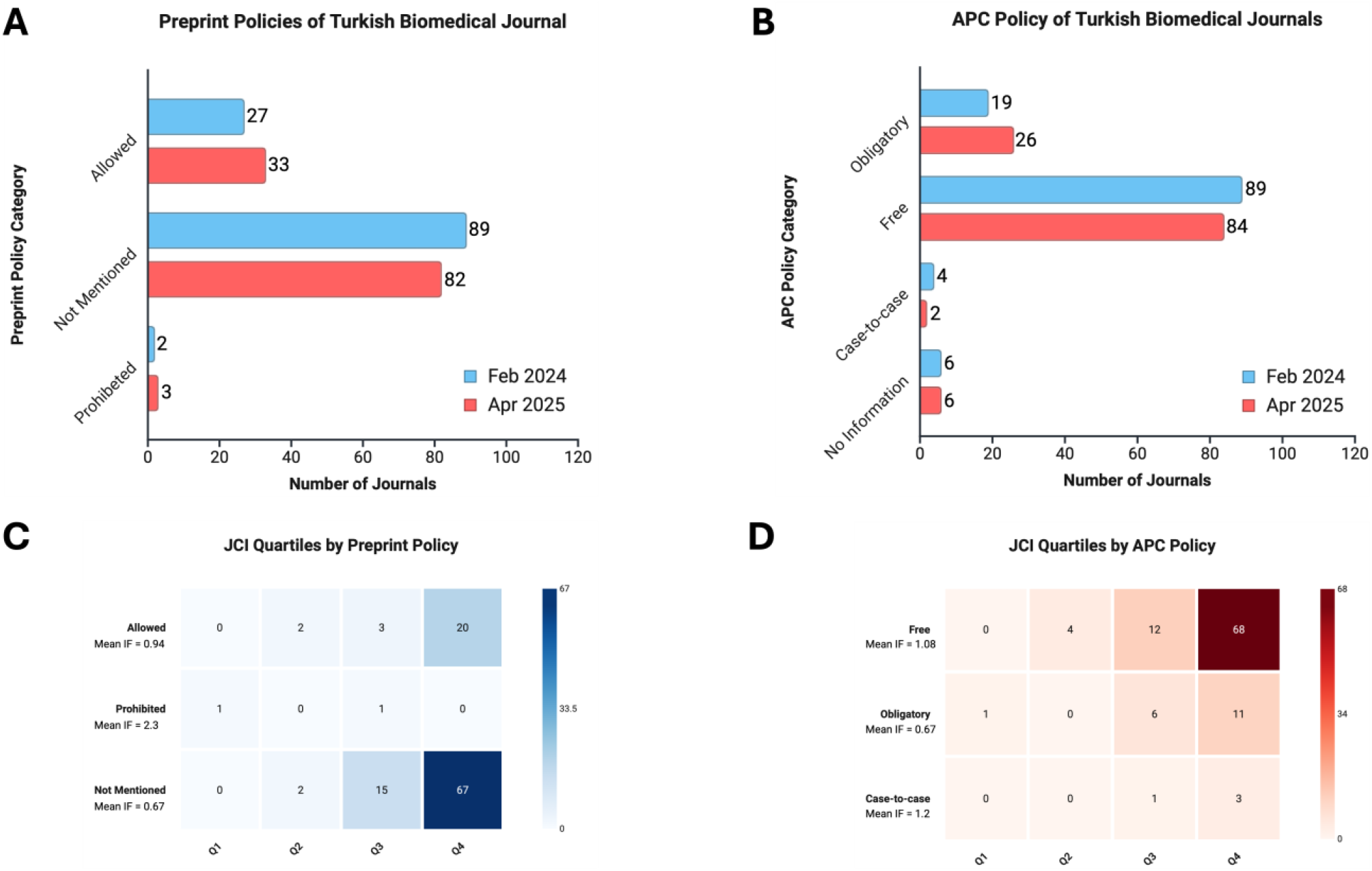
Preprint policy and article processing charge (APC) breakdown of Turkish biomedical journals. **(A)** Change in declared preprint policies of Turkish biomedical journals between February 2024 and April 2025. Journals that explicitly allowed preprints increased from 27 to 33, while those with no policy slightly decreased. **(B)** Change in APC policies of the same journals over the same period. A small rise in journals requiring obligatory APCs was observed. **(C)** Heatmap showing the distribution of journals across JCI quartiles by preprint policy, alongside mean Journal Impact Factor (IF). Journals that allow preprints are more concentrated in higher quartiles compared to those that do not mention or prohibit them. **(D)** Heatmap showing JCI quartile distribution by APC policy. Journals with case-to-case or obligatory APCs are associated with higher mean IFs and more frequent placement in Q1 and Q2.

The implications of these policy differences are further reflected in journal performance metrics. As shown in the heatmap (**Figure 1C**), journals that allow preprints are more likely to appear in higher JCI quartiles (especially Q1 and Q2) and have a higher mean impact factor (0.94) compared to those that prohibit (2.3, though based on very few cases) or do not mention preprints (0.67). While these findings do not suggest causation, they underscore a potential link between preprint openness and higher-performing journals.

### 1.2. Article Processing Charge Policies

We also examined APC policies, grouping them into:

- **Obligatory**: Journals that always charge a fee for publication.
- **Free**: No publication charges to authors.
- **Case-to-case**: Charges apply only under certain conditions (e.g., article type, page length).
- **No Information**: No public declaration of APC policy found.

Between the two assessment periods, we noted a slight increase in journals with obligatory APCs (from 19 to 26), accompanied by a minor decrease in “free” journals (from 89 to 84). This indicates that APCs are becoming more common among Turkish biomedical journals, potentially impacting submission decisions, especially for early-career or unfunded researchers (**Figure 1B**).

When examining journal performance relative to APC policy, journals with case-to-case or obligatory APC models tended to occupy higher JCI quartiles and exhibited higher mean impact factors (1.2 and 1.08, respectively), compared to free journals (mean IF = 0.67) (**Figure 1D**). This suggests that journals with structured APC policies may be more established or competitive in the academic publishing ecosystem.

### 2. Participant Demographics

A total of 103 medical academics participated in the study. Among those who reported sex (n = 98), 51% were female and 49% were male. Age (M = 39.56, SD = 12.64) distribution ranged from early 20s to over 70, with a wide representation from junior researchers to senior professors. In terms of academic titles, professors (28.1%) and residents (29.2%) made up the largest groups (**Table 1**).

**Table 1.**
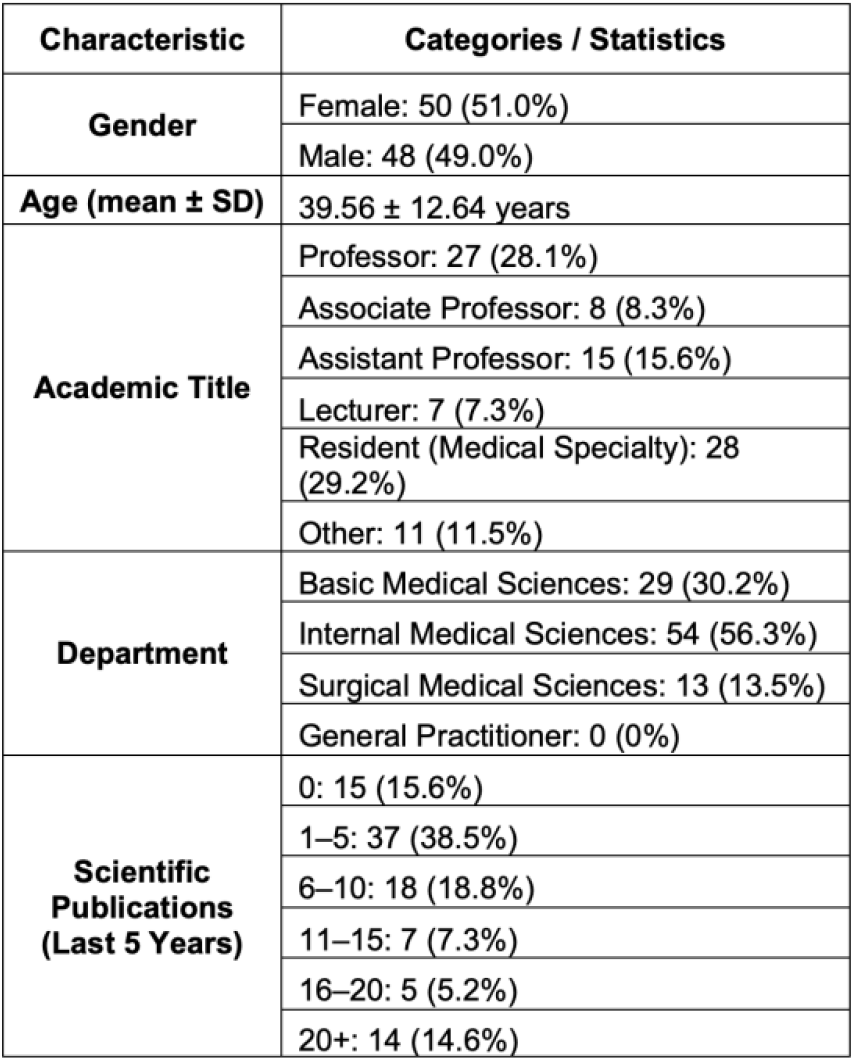
Demography of the participants.

| Characteristic | Categories / Statistics |
| --- | --- |
| Gender | Female: 50 (51.0%) |
|  | Male: 48 (49.0%) |
| Age (mean ± SD) | 39.56 ± 12.64 years |
| Academic Title | Professor: 27 (28.1%) |
|  | Associate Professor: 8 (8.3%) |
|  | Assistant Professor: 15 (15.6%) |
|  | Lecturer: 7 (7.3%) |
|  | Resident (Medical Specialty): 28 (29.2%) |
|  | Other: 11 (11.5%) |
| Department | Basic Medical Sciences: 29 (30.2%) |
|  | Internal Medical Sciences: 54 (56.3%) |
|  | Surgical Medical Sciences: 13 (13.5%) |
|  | General Practitioner: 0 (0%) |
| Scientific Publications (Last 5 Years) | 0: 15 (15.6%) |
|  | 1–5: 37 (38.5%) |
|  | 6–10: 18 (18.8%) |
|  | 11–15: 7 (7.3%) |
|  | 16–20: 5 (5.2%) |
|  | 20+: 14 (14.6%) |

### 3. Awareness of Preprints

A substantial portion of participants lacked awareness of preprints. Only 42.9% of respondents reported familiarity with the term, while 21.4% had never heard of it (**Figure 2A**).

**Figure 2.**
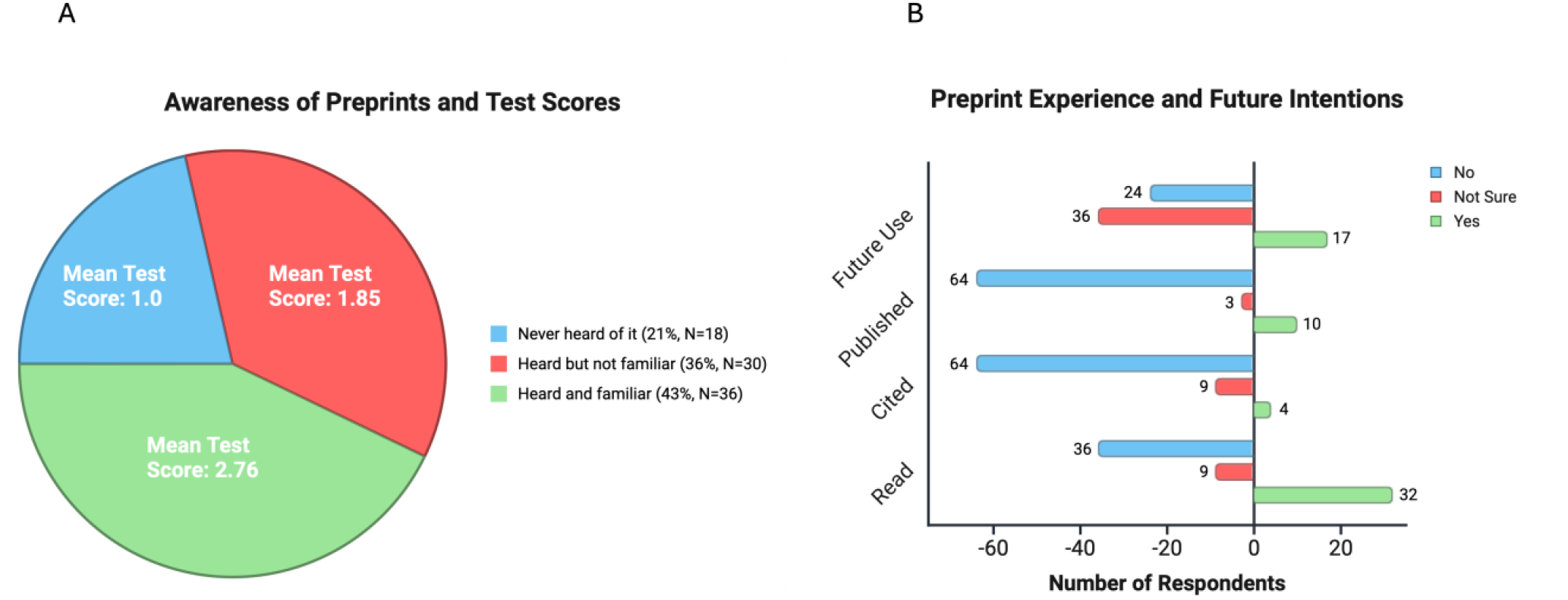
Awareness, knowledge, and experience with preprints among medical academics. **(A)** Pie chart illustrating participants’ awareness of preprints, categorized as “Never heard of it,” “Heard but not familiar,” and “Heard and familiar.” Overlaid values indicate the mean knowledge test scores (range: 0–4) for each awareness group, revealing a clear gradient between awareness and preprint literacy. **(B)** Horizontal bar chart showing the distribution of responses regarding participants’ preprint-related behaviours and intentions, including whether they have read, cited, published, or plan future use of preprints. Responses are grouped by “Yes,” “No,” and “Not sure,” highlighting overall low levels of direct engagement with preprints.

### 4. Knowledge of Preprints

To quantify participants’ objective knowledge of preprints, a “Preprint Test Score” was calculated based on responses to four multiple-choice questions. Participants received one point for each correct answer, resulting in a total score ranging from 0 to 4.

Overall, the mean Preprint Test Score across all respondents was moderate (M = 2.07), with variation seen across both age groups and academic disciplines. Participants who reported being familiar with the term “preprint” scored highest (M = 2.76), while those who had never heard of the term scored the lowest (M = 1.00), suggesting internal validity of the test score (**Figure 2A**).

### 5. Experience with Preprints

Engagement with preprints was low across all indicators. Only 10 participants had published on a preprint server, 32 had read one, and just 4 had cited a preprint in their scientific writing (**Figure 2B**).

### 6. Attitudes Toward Future Use

When asked whether they would consider publishing a future manuscript as a preprint, only 22.1% said yes, while 46.7% were unsure and 31.2% said no (**Figure 2B**).

### 7. Subgroup Analyses

### 7.1. Subgroup analysis by age

Participants were categorized into two age groups: younger (<40 years, n=40) and older (≥40 years, n=29). The mean ages were 30.85 and 51.86 years, respectively. The older group demonstrated greater familiarity with preprints and higher preprint test scores (M = 2.20, SD = 1.31 vs. M = 1.97, SD = 1.60). While only 2.5% of younger participants had published a preprint, 24.1% of older participants had done so. Further, younger participants showed less openness to future use, with 17.5% considering preprint publication compared to 27.6% in the older group, and a larger proportion remaining undecided (62.5%).

Interestingly, younger participants expressed more favorable attitudes toward the role of preprints in scientific development and reforming the peer review system, whereas older participants valued conventional indicators of study quality such as citation counts and journal prestige. These differences are illustrated in **Table 2**.

**Table 2.**
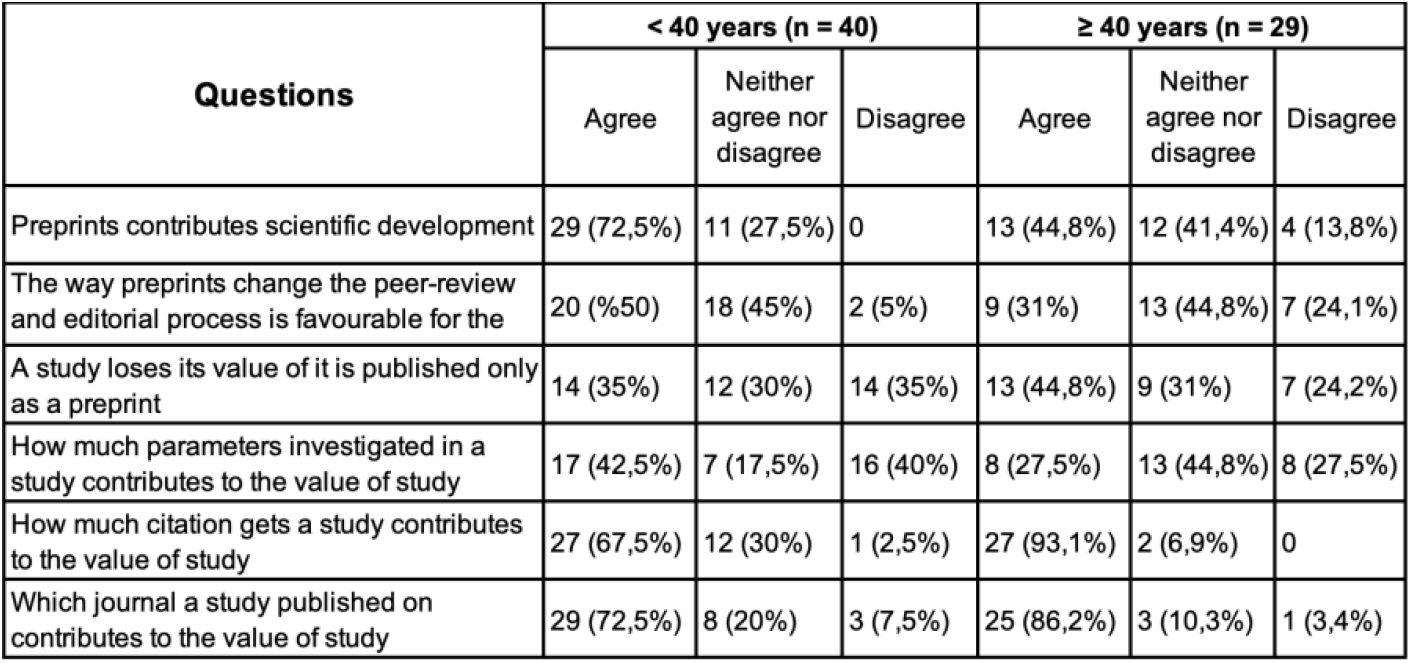
Attitudes and perceptions toward preprints by age group.

| Questions | < 40 years (n = 40) |  |  | ≥ 40 years (n = 29) |  |  |
| --- | --- | --- | --- | --- | --- | --- |
|  | Agree | Neither agree nor disagree | Disagree | Agree | Neither agree nor disagree | Disagree |
| Preprints contributes scientific development | 29 (72,5%) | 11 (27,5%) | 0 | 13 (44,8%) | 12 (41,4%) | 4 (13,8%) |
| The way preprints change the peer-review and editorial process is favourable for the | 20 (50%) | 18 (45%) | 2 (5%) | 9 (31%) | 13 (44,8%) | 7 (24,1%) |
| A study loses its value if it is published only as a preprint | 14 (35%) | 12 (30%) | 14 (35%) | 13 (44,8%) | 9 (31%) | 7 (24,2%) |
| How much parameters investigated in a study contributes to the value of study | 17 (42,5%) | 7 (17,5%) | 16 (40%) | 8 (27,5%) | 13 (44,8%) | 8 (27,5%) |
| How much citation gets a study contributes to the value of study | 27 (67,5%) | 12 (30%) | 1 (2,5%) | 27 (93,1%) | 2 (6,9%) | 0 |
| Which journal a study published on contributes to the value of study | 29 (72,5%) | 8 (20%) | 3 (7,5%) | 25 (86,2%) | 3 (10,3%) | 1 (3,4%) |

### 7.2. Subgroup analysis by academic discipline

Participants were also grouped by academic department into basic sciences (n=24) and clinical sciences (n=45). The mean ages were 37.63 and 40.78 years, respectively. Preprint test scores and past use were similar across both groups. However, future intentions diverged: 29.2% of basic science participants considered future preprint use, compared to 17.8% in the clinical sciences group. Additionally, 37.8% of clinical faculty reported not considering preprint use in the future, suggesting a more cautious stance.

Attitudinal data further revealed that clinical scientists were more goal-oriented and focused on applicability in practice, whereas basic scientists placed more emphasis on the breadth of research parameters and openness to publication reform. Notably, more basic science participants believed that studies published only as preprints may lack value (58.4%) compared to clinical scientists (28.9%). These findings are visualized in **Table 3**.

**Table 3.** Attitudes and perceptions toward preprints by academic discipline.

| Questions | Basic Sciences (n = 24) |  |  | Clinical Sciences (n = 45) |  |  |
| --- | --- | --- | --- | --- | --- | --- |
|  | Agree | Neither agree nor disagree | Disagree | Agree | Neither agree nor disagree | Disagree |
| Preprints contributes scientific development | 16 (66,6%) | 8 (33,4%) | 0 | 26 (57,8%) | 15 (33,3%) | 4 (8,9%) |
| The way preprints change the peer-review and editorial process is favourable for the future of science | 13 (54,2%) | 10 (41,7%) | 1 (4,2%) | 16 (35,5%) | 21 (46,7%) | 8 (17,8%) |
| A study loses its value if it is published only as a preprint | 14 (58,4%) | 4 (16,7%) | 6 (25%) | 13 (28,9%) | 17 (37,8%) | 4 (8,9%) |
| How much parameters investigated in a study contributes to the value of study | 13 (54,2%) | 7 (29,2%) | 4 (16,6%) | 12 (26,7%) | 13 (28,9%) | 18 (44,4%) |
| How much citation gets a study contributes to the value of study | 17 (70,9%) | 6 (25%) | 1 (4,2%) | 37 (82,2%) | 8 (17,8%) | 0 |
| Which journal a study published on contributes to the value of study | 16 (66,6%) | 7 (29,2%) | 1 (4,2%) | 38 (84,4%) | 4 (8,9%) | 3 (6,6%) |

### 8. Barriers to Preprint Adoption

Understanding why academics and editors hesitate to engage with preprints is critical for developing targeted interventions. In this study, qualitative responses from both survey participants and journal editors were analyzed thematically to uncover common concerns.

#### 8.1. Perceived Barriers Among Medical Academics (Survey Respondents)

Thematic analysis of open-ended responses revealed common concerns including fear of plagiarism or idea theft, lack of academic recognition, and insufficient knowledge about preprints. These barriers are summarized in **Table 4**.

**Table 4.**
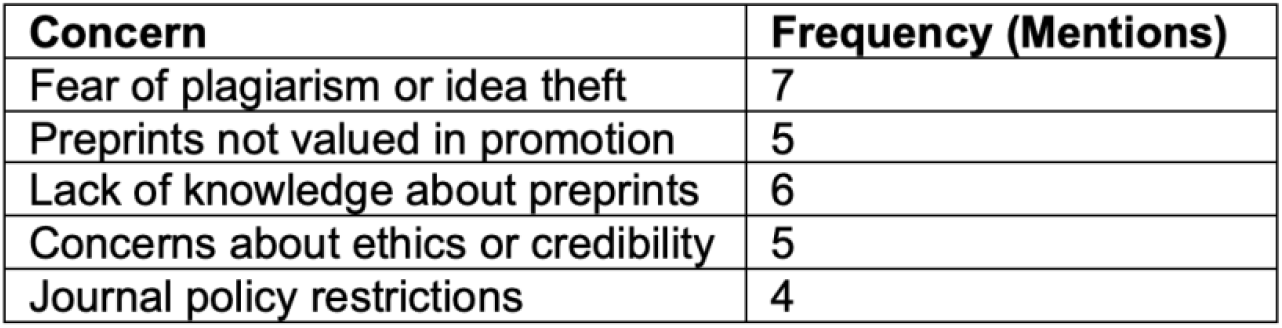
Common barriers to preprint use identified in participant comments.

| Concern | Frequency (Mentions) |
| --- | --- |
| Fear of plagiarism or idea theft | 7 |
| Preprints not valued in promotion | 5 |
| Lack of knowledge about preprints | 6 |
| Concerns about ethics or credibility | 5 |
| Journal policy restrictions | 4 |

Participants were asked an open-ended question regarding their personal concerns or hesitations about using preprints. Thematic analysis revealed several recurring barriers:

- **Fear of plagiarism or idea theft:** A frequently mentioned concern was the potential for unreviewed ideas to be copied or republished without attribution. This concern appeared especially prominent among early-career researchers.
- **Preprints not valued in promotion:** Several participants indicated that preprints are not acknowledged in institutional promotion or academic evaluation processes. As a result, preprints were seen as a risky or unrewarding form of dissemination.
- **Lack of knowledge about preprints:** Many respondents were unsure about how to submit preprints, what platforms were reputable, or how preprints interact with formal journal submissions.
- **Concerns about ethics or credibility:** Some participants questioned whether preprints, by not undergoing peer review, could contribute to the spread of low-quality or misleading research.
- **Journal policy restrictions:** A few respondents mentioned that they avoided preprints because they believed many journals would reject submissions previously shared as preprints, even if that was not explicitly stated.

These findings suggest that barriers are shaped by both institutional norms and practical uncertainties.

#### 8.2. Editorial Perspectives from Turkish Biomedical Journals

Responses from seven journal editors in Türkiye revealed a spectrum of attitudes toward preprints, ranging from cautious support to open opposition.

One editor expressed support for preprints as a tool to promote transparency and protect authorship in a landscape where idea theft is perceived to be common. Some other editors described preprints as unnecessary for their journal, noting that they already publish accepted articles promptly and that their infrastructure does not support additional processing. Another editor viewed preprints sceptically due to the possibility of their misuse and the ethical complexity of assigning multiple DOIs to similar content. Another editorial opinion highlighted concerns over duplicate publication and the risk that preprints with assigned DOIs might be flagged as plagiarism in similarity checks. Despite these concerns, one editor predicted that preprints would become more widely accepted in the future, potentially reshaping the landscape of academic publishing in Türkiye.

Overall, the responses reflected uncertainty, infrastructural limitations, and a lack of standardization—factors that likely influence journal-level policies and affect how academics perceive the safety and legitimacy of preprint publishing.

## DISCUSSION

Our findings reveal a notable gap in both awareness and practical engagement with preprints among medical academics at a major university in Istanbul. Despite the global momentum toward open science and rapid communication (13), many respondents exhibited limited familiarity with the concept, and a majority expressed hesitance or scepticism about its use. A recent global survey found that approximately 10% of medical and health sciences researchers in the United States were unfamiliar with preprints, and around 40% had never posted one (14). In contrast, one-fifth of our participants had never heard of preprints, and four-fifths had never submitted one, highlighting a relatively greater degree of unawareness and hesitancy in our sample.

Misconceptions regarding peer review, duplicate publication, and ethical validity were widespread among respondents, underscoring the need for targeted education and clearer guidance. Maggio et al. have proposed formally integrating preprint education into health professions curricula to improve understanding and normalize early dissemination practices (15). Moreover, research shows that students and early career researchers often struggle to distinguish preprints from peer-reviewed journal articles, reflecting a significant knowledge gap (16). Consistent with these findings, our participants under the age of 40 demonstrated lower preprint knowledge, emphasizing the urgency of tailored interventions for this group.

Journal editors’ responses reflected a similar ambivalence. While some recognized the potential of preprints for visibility and transparency, others raised concerns about incompatibility with traditional editorial workflows, ethical ambiguity, and the potential for misuse. Despite such scepticism, studies have shown that peer-reviewed articles that were first posted as preprints tend to receive more citations and broader attention (17), suggesting tangible benefits to early sharing.

Our complementary policy analysis of 118 Turkish biomedical journals further highlights the cultural and structural barriers impeding broader preprint adoption. Only a small proportion of journals explicitly permitted preprints, while the vast majority either prohibited them or failed to mention them altogether. This lack of clear guidance stands in contrast to many high-ranking international clinical journals, which now explicitly allow or even encourage the submission of manuscripts previously posted as preprints (18). The absence of formal policies among Turkish journals likely contributes to hesitation among authors, reinforcing an environment of academic conservatism and uncertainty—a trend previously noted in the literature (8).

However, our longitudinal review of journal policies between February 2024 and April 2025 suggests a slow but positive shift in this landscape. During this period, six additional journals formally adopted policies allowing preprints, while only one new prohibition was identified. Although incremental, this trend points toward a growing acceptance and normalization of preprint use within the Turkish biomedical publishing ecosystem. As journal policies become more explicit and aligned with global standards, uncertainty among researchers may diminish, potentially encouraging wider adoption of preprints in academic practice (19).

Additionally, variability in Article Processing Charge (APC) policies—particularly the prevalence of “free” and “case-to-case” models—may influence authors’ motivations. While one of the advantages of preprints is their cost-free accessibility, this incentive may be undercut if authors are already publishing in fee-free journals or are unaware of preprint benefits (20). Financial considerations, combined with policy ambiguity, could thus create a disincentive for broader adoption.

Our subgroup analyses revealed how age and academic discipline influence perceptions of preprints. Older academics (≥40 years) were more likely to have published on preprint servers and scored higher on objective knowledge measures, but also displayed more conservative attitudes, valuing journal prestige and citation counts. Younger academics (<40 years), on the other hand, were more pessimistic about the future potential of preprints, despite their limited experience. These generational differences may reflect an evolving academic culture that increasingly values transparency, speed, and accessibility.

This interpretation aligns with the findings of Fraser et al., who noted that junior researchers often use preprints to increase the visibility of their work, whereas senior researchers are more motivated by competitive concerns such as staking a priority claim (20).

An interesting finding is that, even though younger academics display reformist attitudes towards conventional publishing process, they show less openness towards use of preprints in future. It can be due to their inexperienced status or misconceptions regarding preprint, which can be validated by their lower preprint test score.

We also observed differences between basic and clinical science faculty. While overall knowledge levels were similar, clinical faculty were more hesitant about preprint use. This may be due to the conservative nature of clinical research, which is closely tied to patient safety, regulatory compliance, and reliance on peer-reviewed evidence. As previously described, clinical medicine tends to be more cautious regarding the role of preprints in academic communication (21).

It is important to acknowledge that this study was conducted at a single academic institution in İstanbul. As such, our findings represent a localized snapshot and should be interpreted with caution. Broader, multi-centre studies will be necessary to determine whether these patterns hold across other regions and institutions in Türkiye.

To enable broader and more equitable adoption of preprints in the country, we recommend:

- Integration of preprints into academic evaluation and promotion criteria,
- Development of clear and accessible editorial policies that explicitly state preprint compatibility,
- Cross-disciplinary and inter-institutional dialogue to address differing perceptions of scientific quality and publishing ethics,
- Encouragement of national or regional preprint platforms tailored to local academic contexts.
- Bridging both the awareness and policy gaps—and accommodating the diversity of views within academic institutions—is essential for aligning Türkiye’s scientific publishing culture with international standards.

## CONCLUSION

This study reveals a substantial gap in both awareness and engagement with preprints among medical academics at a prominent university in İstanbul. Despite increasing international recognition of preprints as a valuable tool for early research dissemination, many participants demonstrated limited familiarity and held misconceptions regarding their ethical standing, academic value, and compatibility with traditional publishing norms.

Our analysis of Turkish biomedical journal policies offers important context for this hesitancy. Most journals either prohibit preprints or provide no clear guidance, creating a policy vacuum that likely discourages uptake and reinforces uncertainty among potential preprint authors. However, longitudinal data from our analysis indicate that this landscape may be gradually shifting— between February 2024 and April 2025, several journals adopted more permissive policies regarding preprints, suggesting a slow but positive trend toward greater acceptance within Türkiye’s biomedical publishing ecosystem.

Subgroup analyses further emphasized the diversity of perspectives within the academic community. Preprint attitudes were strongly influenced by age and academic discipline: older academics and clinical scientists were more likely to hold traditional views, while basic scientists expressed more openness to evolving publishing practices. Exceptionally, younger scientists were not sure about future use of preprints while displaying reformist attitudes.

While these findings are not generalizable to the entire Turkish medical academic landscape, they provide valuable insight into the institutional, cultural, and structural factors influencing preprint adoption. Strategic, context-sensitive interventions—including targeted education, transparent and consistent journal policies, and institutional recognition of preprints—are essential to support a more open, timely, and inclusive scientific communication ecosystem.

## Data Availability

All data produced in the present study are available upon reasonable request to the authors

## ACKNOWLEDGEMENT

We would like to thank Burak Kızılca for the assistance provided during collection of journal policies. All graphs Created in https://BioRender.com

## AUTHOR CONTRIBUTIONS

Conceptualization, methodology, writing—original draft preparation, writing—review and editing M.S., B.K. and Z.A. All authors have read and agreed to the published version of the manuscript.

## FUNDING

This study did not receive any funding.

## ETHICAL APPROVAL

The study was approved by the Marmara University Faculty of Medicine Non-Drug and Medical Device Research Ethics Committee (protocol code 09.2024.600 and date of 17 May 2024)

## CONFLICTS OF INTEREST

The authors declare no conflict of interest.

## Notes

### Competing Interest Statement

The authors have declared no competing interest.

